# Using an electronic frailty index to predict adverse outcomes in geriatric COVID-19 patients: data from the Stockholm GeroCovid study

**DOI:** 10.1101/2021.12.03.21267254

**Authors:** Jonathan K. L. Mak, Maria Eriksdotter, Martin Annetorp, Ralf Kuja-Halkola, Laura Kananen, Anne-Marie Boström, Miia Kivipelto, Carina Metzner, Viktoria Bäck Jerlardtz, Malin Engström, Peter Johnson, Lars Göran Lundberg, Elisabet Åkesson, Carina Sühl Öberg, Maria Olsson, Tommy Cederholm, Sara Hägg, Dorota Religa, Juulia Jylhävä

**Author notes:** Shared last authorship (Dorota Religa, Juulia Jylhävä). Correspondence: Dorota Religa, Department of Neurobiology, Care Sciences and Society, Karolinska Institutet, Blickagången 16, 141 52 Huddinge, Sweden.

## Abstract

**Background:** The Clinical Frailty Scale (CFS) is a strong predictor for worse outcomes in geriatric COVID-19 patients, but it is less clear whether an electronic frailty index (eFI) constructed from routinely collected electronic health records (EHRs) provides similar predictive value. This study aimed to investigate the predictive ability of an eFI in comparison to other frailty and comorbidity measures, using mortality, readmission, and the length of stay as outcomes in geriatric COVID-19 patients.

**Methods:** We conducted a retrospective cohort study using EHRs from nine geriatric clinics in Stockholm, Sweden, comprising 3,405 COVID-19 patients (mean age 81.9 years) between 1/3/2020 and 31/10/2021. Frailty was assessed using a 48-item eFI developed for Swedish geriatric patients, the CFS, and Hospital Frailty Risk Score (HFRS). Comorbidity was measured using the Charlson Comorbidity Index (CCI). We analyzed in-hospital mortality and 30-day readmission using logistic regression and area under receiver operating characteristic curve (AUC). 30-day and 6-month mortality were modelled by Cox regression, and the length of stay by linear regression.

**Results:** Controlling for age and sex, a 10% increase in the eFI was associated with higher risks of in-hospital mortality (odds ratio [OR]=2.84; 95% confidence interval=2.31-3.51), 30-day mortality (hazard ratio [HR]=2.30; 1.99-2.65), 6-month mortality (HR=2.33; 2.07-2.62), 30-day readmission (OR=1.34; 1.06-1.68), and longer length of stay (*β*=2.28; 1.90-2.66).

The CFS, HFRS and CCI similarly predicted these outcomes, but the eFI had the best predictive accuracy for in-hospital mortality (AUC=0.775).

**Conclusions:** An eFI based on routinely collected EHRs can be applied in identifying high-risk geriatric COVID-19 patients.

## INTRODUCTION

Accumulating evidence has demonstrated that frailty, an ageing-related state of heightened vulnerability to stressors, is a strong predictor of COVID-19 mortality in hospitalized patients, providing predictive value beyond chronological age (1–6). Most of the studies measured frailty using the Clinical Frailty Scale (CFS) (7), a clinical judgement-based tool with generally high accuracy, but it requires in-person evaluation and may not always be available. An alternative clinical measure of frailty is the Hospital Frailty Risk Score (HFRS) (8), calculated based on *International Classification of Diseases, Tenth Revision* (ICD-10) codes. However, only some studies (9), but not others (1,10,11), have found an association between the HFRS and COVID-19 mortality in hospitalized patients. The frailty index (FI) is one of the best-validated frailty measures (12), but its assessment is resource-intensive, and only few studies with relatively small sample sizes have examined its predictive value for COVID-19 mortality (13,14).

We recently developed an electronic frailty index (eFI) based on routinely collected electronic health records (EHRs) for geriatric patients in the Stockholm region, Sweden and showed that it has a good predictive ability for adverse outcomes in non-COVID-19 patients (15). We based the Swedish eFI model on the US model by Pajewski *et al*. (16), as it was highly compatible with data contained in the hospital EHRs in the Stockholm region. Given the ongoing COVID-19 pandemic and its burden on healthcare, it is important to investigate whether an automated eFI could be used to identify high-risk COVID-19 patients, guide decision making and aid in planning of follow up care. Therefore, this study aimed to assess the predictive value of the eFI, in comparison to other validated frailty and comorbidity measures, for mortality, readmission, and the length of stay in older COVID-19 patients.

## METHODS

### Study population

We extracted EHRs of patients from nine geriatric clinics in the Stockholm region between March 1, 2020 and October 31, 2021. Only patients treated for COVID-19 were included in the present analyses; other patients admitted to the same clinics during the study period were analyzed in a separate paper (15). COVID-19 was determined by either a positive result of reverse transcriptase polymerase chain reaction (RT-PCR), or a clinical diagnosis for those with a negative RT-PCR but with typical symptoms and computed tomography scan findings (ICD-10 codes U07.1 or U07.2; n=3,272). We also included those with a medical history or post-infectious condition of COVID-19 (ICD-10 codes U08.9, U09.9, or U10.9; n=133). Exclusion criteria were: (i) missing discharge information, (ii) hospital stay <24h, and (iii) insufficient data for calculation of the eFI. For patients with multiple admissions, only the first admission with a COVID-19 diagnosis was analyzed.

Given the large fluctuation of COVID-19 infection rates and mortality rates during the pandemic in Sweden, we considered four study periods according to the statistics of COVID-19 cases and vaccination coverage by the Public Health Agency of Sweden (17): period 1 (“1^st^ wave”) was defined as March 1, 2020 to August 31, 2020; period 2 (“2^nd^ wave”) as September 1, 2020 to January 31, 2021; period 3 (“3^rd^ wave”) as February 1, 2021 to April 20, 2021; and period 4 (the time after which most of the patients in our sample were assumed to be fully vaccinated) as April 21, 2021 to October 31, 2021. Number of admitted geriatric COVID-19 patients throughout the four periods is shown in **Supplementary Figure 1**.

This study was approved by the Swedish Ethical Review Authority (Dnr 2020-02146, 2020-03345, 2021-00595, 2021-02096).

### Exposures

Three frailty measures (eFI, CFS, HFRS) and a comorbidity measure (Charlson Comorbidity Index [CCI]) were used:

- An eFI was derived based on a US model by Pajewski *et al*. (16). Details on its development and coding of the eFI items are described elsewhere (15). Briefly, a list of 48 deficits from disease diagnoses, functional abilities and other health indicators such as oral health, falls and weight loss, and laboratory measures were included in the eFI (**Supplementary Table 1**), following the principles of the deficit accumulation model (12,18). Those with <30 non-missing items, or those missing more than half of the functioning and/or laboratory deficits were excluded from the analysis. Patients were categorized into four eFI groups: fit (≤0.15), mild frailty (>0.15–0.2), moderate frailty (>0.2–0.25), and severe frailty (>0.25), analogously as described by Clegg *et al*. (18).
- The CFS was scored by a physician or trained nurse at admission, ranging from 1 (“very fit”) to 9 (“terminally ill”), and was categorized into three groups: 1–3, 4–5, and 6–9.
- The HFRS was calculated based on 109 frailty-associated ICD-10 codes, using the algorithm described by Gilbert *et al*. (8). Patients were grouped into low (<5), intermediate (5–15) and high (>15) risk of frailty.
- The CCI was computed based on ICD-10 codes, following an algorithm adapted for Swedish settings (19). It was used as a continuous variable.

### Outcomes

Study outcomes included (i) in-hospital mortality, (ii) 30-day mortality, (iii) 6-month mortality, (iv) 30-day readmission to any of the nine geriatric clinics included in this study and considering individuals who were discharged to home after the first admission, and (v) the length of hospital stay. The outcomes were assessed based on dates of admission, discharge, and vital status extracted from EHRs.

### Analysis

We calculated odds ratios (ORs) and 95% confidence intervals (CIs) for in-hospital mortality and 30-day readmission using logistic regression. Area under the receiver operating characteristic curve (AUC) was used to assess the predictive accuracy of the eFI and other measures of frailty and comorbidity. Cox models were used to estimate the hazard ratios (HRs) for 30-day and 6-month mortality, with the associated Harrell’s C-statistics for predictive accuracy. The 95% CIs for the Harrell’s C-statistics were calculated using 1,000 bootstrapping resampling. Linear regression was used for the length of stay. Age and sex were adjusted for in all models. We also performed subgroup analyses for the eFI and in-hospital mortality by the study periods, admitting clinics, age group, and sex. R version 4.0.5 was used for all analyses.

## RESULTS

Of the 3,405 COVID-19 geriatric patients, 53.8% were women and the mean age was 81.9 years (**Table 1**). In-hospital mortality rate was 9.5%, and the average length of stay was 8.8 days. The median eFI was 0.182 (interquartile range=0.141–0.227; range=0–0.396), with 13.5% classified as severely frail; there was no significant sex difference in the frailty scores. Across the waves of the COVID-19 pandemic (**Supplementary Table 2**), those admitted later appeared to be younger, have lower frailty scores, lower in-hospital death rates, and shorter length of stay. The eFI was moderately correlated with other measures, Spearman’s correlations of which were 0.464 with CFS, 0.365 with HFRS, and 0.419 with CCI (**Supplementary Table 3**).

**Table 1.**
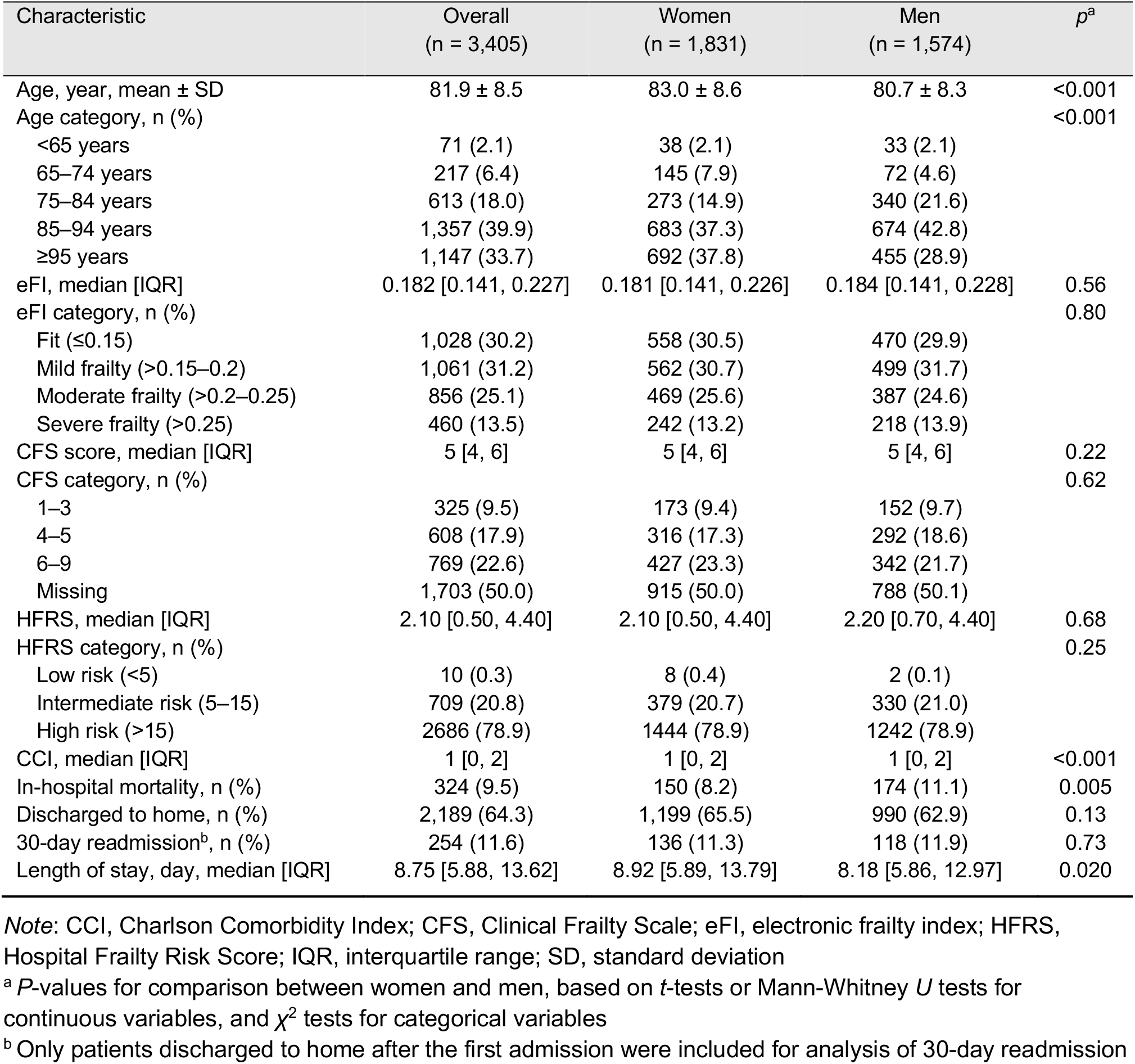
Sample characteristics by sex

**Table 2** shows the associations of frailty and comorbidity with mortality outcomes. The eFI (OR per 10% increase=2.84, 95% CI=2.31–3.51), as well as other measures, were significantly associated with in-hospital mortality after adjusting for age and sex. However, among all measures, adding the eFI to a model with age and sex yielded the greatest AUC of 0.775 (**Figure 1A**). Considering the potential heterogeneity of patients admitted during different time periods and to different clinics, we performed a subgroup analysis and found that the eFI had a similar predictive accuracy for in-hospital mortality across the four time periods (AUCs 0.729–0.788) and the nine clinics (AUCs 0.740–0.805) (**Supplementary Figure 2**). The association between the eFI and in-hospital mortality tended to be stronger in younger age groups (*p*_interaction_=0.005 across the three age groups of <75, 75–84, and ≥85 years) but was similar in men and women (**Supplementary Figure 2**).

**Table 2.** Associations between frailty and comorbidity measures and mortality outcomes (n = 3,405)^a^

| Model | In-hospital mortality | 30-day mortality | 6-month mortality |
| --- | --- | --- | --- |
|  | Adjusted OR (95% CI) | Adjusted HR (95% CI) | Adjusted HR (95% CI) |
| **eFI** |  |  |  |
| Continuous (per 10% increase) | 2.84 (2.31, 3.51)* | 2.30 (1.99, 2.65)* | 2.33 (2.07, 2.62)* |
| Categorical |  |  |  |
| Fit ($\leq 0.15$) | 1 (Ref.) | 1 (Ref.) | 1 (Ref.) |
| Mild frailty ($>0.15-0.2$) | 1.49 (0.98, 2.30) | 1.64 (1.21, 2.23)* | 1.58 (1.24, 2.00)* |
| Moderate frailty ($>0.2-0.25$) | 3.43 (2.33, 5.16)* | 3.18 (2.39, 4.25)* | 2.83 (2.26, 3.55)* |
| Severe frailty ($>0.25$) | 5.44 (3.62, 8.32)* | 4.34 (3.21, 5.88)* | 4.29 (3.38, 5.45)* |
| **CFS (n = 1,702)<sup>b</sup>** |  |  |  |
| Continuous (per point increase) | 1.59 (1.41, 1.82)* | 1.40 (1.28, 1.54)* | 1.42 (1.32, 1.54)* |
| Categorical |  |  |  |
| 1–3 | 1 (Ref.) | 1 (Ref.) | 1 (Ref.) |
| 4–5 | 1.06 (0.56, 2.12) | 1.05 (0.67, 1.66) | 1.28 (0.87, 1.90) |
| 6–9 | 3.41 (1.93, 6.52)* | 2.41 (1.59, 3.66)* | 2.82 (1.95, 4.08)* |
| **HFRS** |  |  |  |
| Continuous (per point increase) | 1.09 (1.05, 1.13)* | 1.07 (1.04, 1.09)* | 1.07 (1.05, 1.09)* |
| Categorical |  |  |  |
| Low risk ($<5$) | 1 (Ref.) | 1 (Ref.) | 1 (Ref.) |
| Intermediate risk (5–15) | 1.66 (1.28, 2.14)* | 1.47 (1.21, 1.77)* | 1.47 (1.26, 1.72)* |
| High risk ($>15$) | No observation | No observation | 0.99 (0.32, 3.08) |
| **CCI** |  |  |  |
| Continuous (per point increase) | 1.22 (1.14, 1.31)* | 1.19 (1.14, 1.25)* | 1.26 (1.22, 1.31)* |

Note: CCI, Charlson Comorbidity Index; CFS, Clinical Frailty Scale; CI, confidence interval; eFI, electronic frailty index; HFRS, Hospital Frailty Risk Score; HR, hazard ratio; OR, odds ratio
<sup>a</sup> All the listed models were multivariate logistic or Cox regression models adjusted for age and sex. eFI, CFS, and HFRS were used as both continuous and categorical variables in separate models, while CCI was used as continuous variable only.
<sup>b</sup> Sample size was smaller in analysis of CFS due to missing data
\* $p < 0.05$

**Figure 1.**
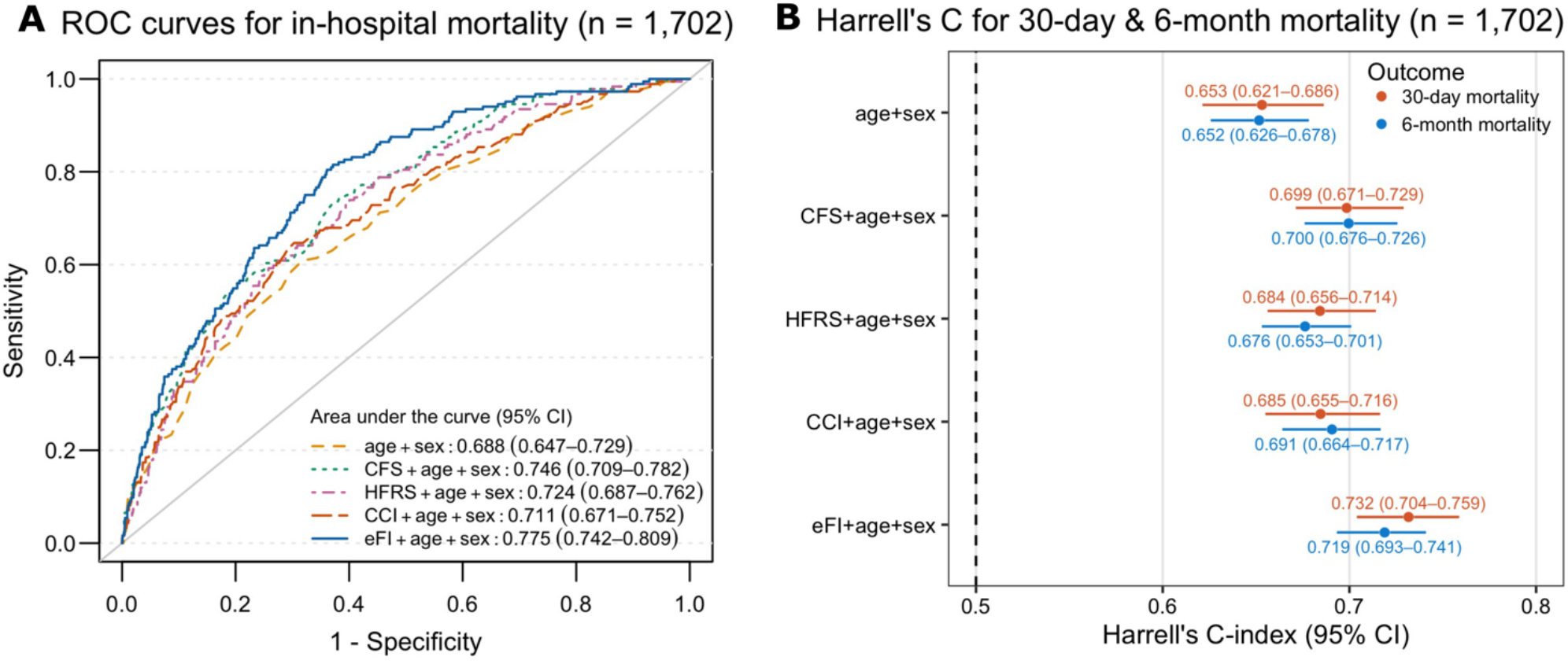
Predictive accuracies of frailty and comorbidity measures for mortality outcomes in patients with all the measures available (n = 1,702). **(A)** Receiver operating characteristics (ROC) curves for in-hospital mortality; **(B)** Harrell’s C-statistics from Cox models for 30-day and 6-month mortality. CFS, HFRS, CCI, and eFI were considered as continuous variables in all the models. Abbreviations: CFS, Clinical Frailty Scale; HFRS, Hospital Frailty Risk Score; CCI, Charlson Comorbidity Index; eFI, electronic frailty index

We followed patients up to six months from admission and observed higher mortality rates among those in the higher eFI categories (**Supplementary Figure 3**). The eFI was associated with 30-day and 6-month mortality, the age- and sex-adjusted HRs per a 10% increase being 2.30 (95% CI=1.99–2.65) and 2.33 (95% CI=2.07–2.62), respectively (**Table 2**). The eFI, compared to the CFS, HFRS, and CCI, also had the best predictive accuracy for 30-day (Harrell’s C=0.732) and 6-month mortality (Harrell’s C=0.719) (**Figure 1B**).

Furthermore, a 10% increase in the eFI was associated with higher odds of 30-day readmission (OR=1.34, 95% CI=1.06–1.68; AUC=0.627) and longer length of stay (*β*=2.28, 95% CI=1.90–2.66) (**Supplementary Tables 4–5**).

## DISCUSSION

In this sample of geriatric COVID-19 patients in Sweden, frailty was significantly associated with mortality, readmission, and longer length of stay. The EHR-based eFI had the highest predictive accuracy for short- and long-term mortality compared to the CFS, HFRS and CCI. Our results thus give support to the use of the eFI for risk assessment and planning of care in hospitalized COVID-19 patients.

Without a “gold standard”, various frailty assessment tools have been used in the literature depending on the study setting and data availability. Previous studies in COVID-19 patients mostly used the CFS (20), which is a simple, efficient, and accurate clinical tool (7). Nevertheless, it requires in-person assessment and may not always be available (CFS was missing in ∼50% of patients in our sample). Some studies used the HFRS but only weak associations with COVID-19 mortality were found (1,10,11). In contrast, the association between the CFS and COVID-19 mortality seems robust (1–6). This may be explained by the slightly dissimilar concepts captured: the CFS focuses mainly on functioning and disability (7), while the diagnosis code-based HFRS is more similar to a comorbidity measure (8). We constructed the eFI based on the deficit accumulation model (12,18), including comorbidities, disabilities, cognition, and laboratory deficits, adhering to the multidimensional definition of frailty. The eFI had moderate correlations with the CFS, HFRS, and CCI, and had good predictive accuracy for in-hospital mortality, suggesting that the multidimensional construct of frailty may predict COVID-19 mortality better than mere disability and multimorbidity. In line with a previous study (10), we found stronger associations between the eFI and in-hospital mortality in younger-old (aged <75) than the oldest-old patients (aged ≥85), indicating a greater relative risk conferred by frailty at younger ages. This finding suggests that the effect of frailty on COVID-19 outcomes is not restricted to the oldest individuals; rather, it should be considered as a risk factor in younger-old patients, too.

Few studies have investigated outcomes other than in-hospital mortality in COVID-19 patients (20). Contributing to the evidence base, we found that frailty tools, especially the eFI, have a good predictive accuracy for 30-day and 6-month mortality. We also found a significant association of the eFI with 30-day readmission and longer length of stay.

However, all the frailty and comorbidity measures had rather poor predictive accuracies for readmission (AUCs 0.62–0.66), which may partly be due to misclassification of patients who were readmitted to other hospitals than the nine included geriatric clinics. This matter warrants further studies in samples with a higher readmission coverage.

This study included a relatively large sample of COVID-19 patients admitted to nine geriatric clinics in the Stockholm region. Our current results further strengthening the clinical utility of the eFI; we have previously observed that the Swedish eFI has a good predictive accuracy for adverse outcomes in geriatric patients with other diagnoses too (15). However, our findings may not be generalizable to other COVID-19 patients, such as those treated in intensive care unit. Due to a lack of data, we were also unable to adjust for several mortality risk factors (e.g., socioeconomic status, smoking) and COVID-19 vaccination. Nevertheless, most geriatric patients in Stockholm were likely to be fully vaccinated after April 21, 2020 (17), and we observed similar predictive ability of the eFI for in-hospital mortality before and after this date.

In summary, to the best of our knowledge, this is the first study that used an eFI based on routinely collected EHRs to identify high-risk COVID-19 patients. Our findings support the use of an eFI for risk stratification in hospitalized geriatric patients with COVID-19.

## Data Availability

All data produced in the present study are available upon reasonable request to the authors.

## ACKNOWLEDGEMENTS

We would like to thank all patients, caregivers and staff who contributed to this study. This study was accomplished within the context of the Swedish National Graduate School for Competitive Science on Ageing and Health (SWEAH) funded by the Swedish Research Council.

