## Supplementary data for "Using an electronic frailty index to predict adverse outcomes in geriatric COVID-19 patients: data from the Stockholm GeroCovid study"

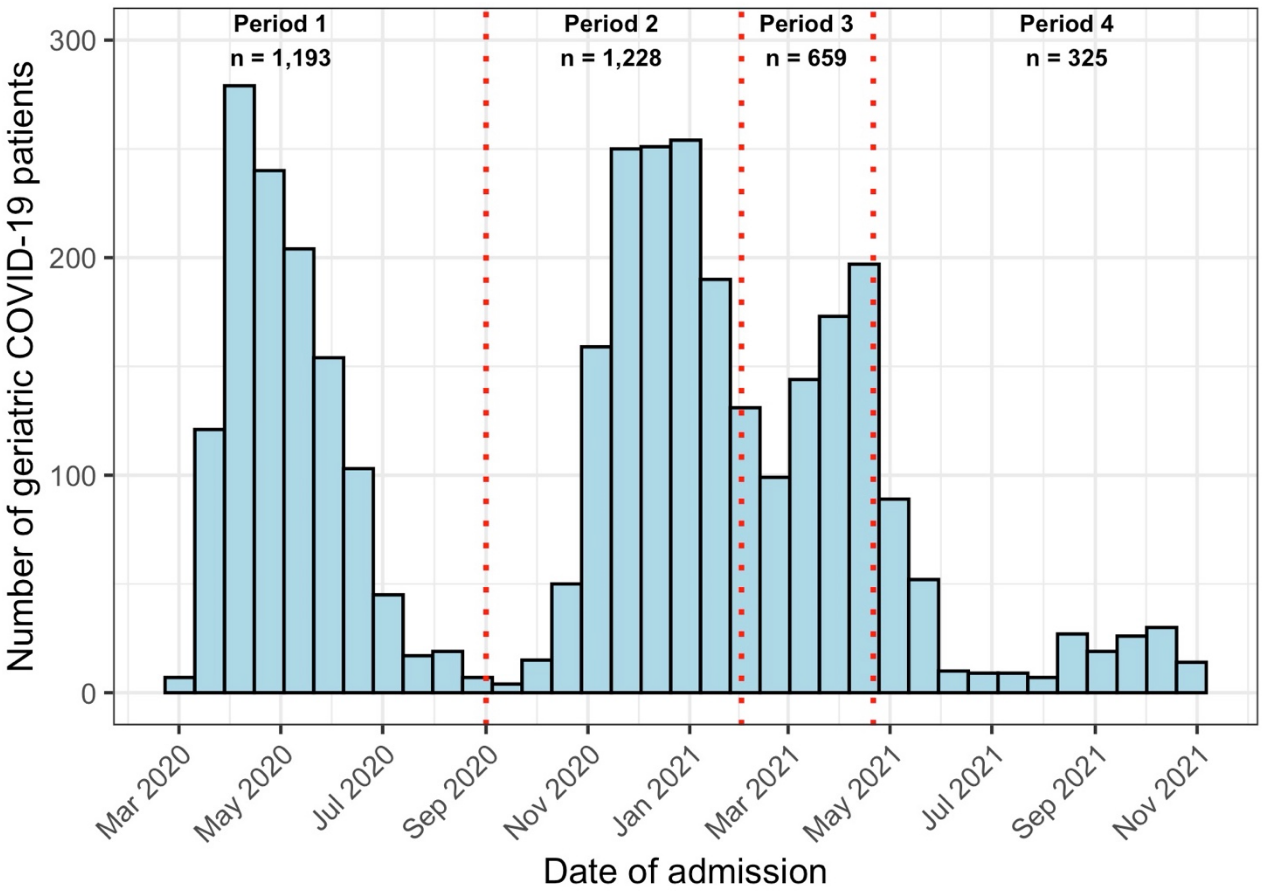

**Supplementary Figure 1.** Number of geriatric COVID-19 patients by date of admission.

Period 1 ("1st pandemic wave") was defined as March 1, 2020 to August 31, 2020; period 2 ("2nd pandemic wave") was defined as September 1, 2020 to January 31, 2021; period 3 ("3rd pandemic wave") was defined as February 1, 2021 to April 20, 2021; period 4 (the time after which most of the patients in our sample were assumed to be fully vaccinated) was defined as April 21, 2021 to October 31, 2021. For patients with multiple admissions, only the first admission with a COVID-19 diagnosis was included in the analysis.

| Subgroup | In-hospital deaths / No. of patients | OR per 10% increase in eFI (95% CI) | AUC |
|---|---|---|---|
| Overall | 324/3,405 | 2.84 (2.31, 3.51) | 0.765 |
| Age | | | |
| <75 years | 17/684 | 5.23 (2.44, 12.00) | 0.801 |
| 75–84 years | 90/1,357 | 3.29 (2.29, 4.78) | 0.727 |
| ≥85 years | 217/1,364 | 2.42 (1.86, 3.17) | 0.691 |
| Sex | | | |
| Women | 150/1,831 | 2.93 (2.18, 3.97) | 0.776 |
| Men | 174/1,574 | 2.75 (2.06, 3.69) | 0.749 |
| Period | | | |
| 1: 3/1/2020 to 31/8/2020 | 168/1,193 | 2.44 (1.80, 3.32) | 0.729 |
| 2: 1/9/2020 to 31/1/2021 | 109/1,228 | 3.94 (2.73, 5.77) | 0.787 |
| 3: 1/2/2021 to 20/4/2021 | 29/659 | 1.68 (0.87, 3.24) | 0.740 |
| 4: 21/4/2021 to 31/10/2021 | 18/325 | 2.31 (1.08, 5.04) | 0.788 |
| Admitting clinics | | | |
| Clinic 1 | 48/421 | 2.22 (1.24, 4.08) | 0.794 |
| Clinic 2 | 15/184 | 1.32 (0.52, 3.48) | 0.740 |
| Clinic 3 | 23/379 | 4.50 (1.97, 10.71) | 0.800 |
| Clinic 4 | 45/362 | 2.83 (1.65, 5.00) | 0.790 |
| Clinic 5 | 34/329 | 3.13 (1.44, 7.13) | 0.805 |
| Clinic 6 | 32/423 | 3.63 (1.88, 7.25) | 0.767 |
| Clinic 7 | 0/30 | No observation | - |
| Clinic 8 | 40/806 | 2.18 (1.25, 3.81) | 0.753 |
| Clinic 9 | 87/471 | 2.82 (1.89, 4.27) | 0.761 |

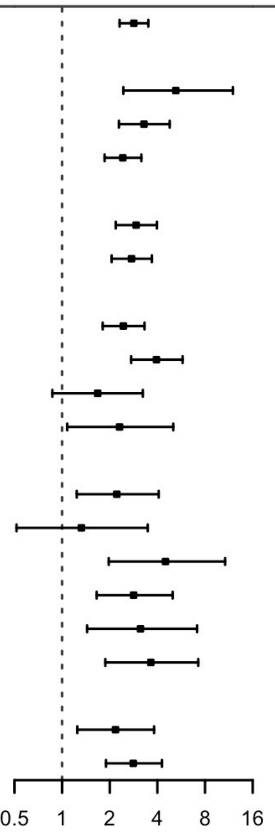

**Supplementary Figure 2.** Subgroup analysis of the associations between the electronic frailty index and in-hospital mortality by age groups, sex, study period, and admitting clinics.

All the listed models were multivariate logistic regression models adjusted for age (continuous) and sex, except when sex was used as the subgroup variable. The four study periods correspond to the 1st pandemic wave, 2nd pandemic wave, 3rd pandemic wave, and the time after which most of the patients in our sample were assumed to be fully vaccinated, respectively. Abbreviations: AUC, area under the receiver operating characteristic curve; CI, confidence interval; OR, odds ratio.

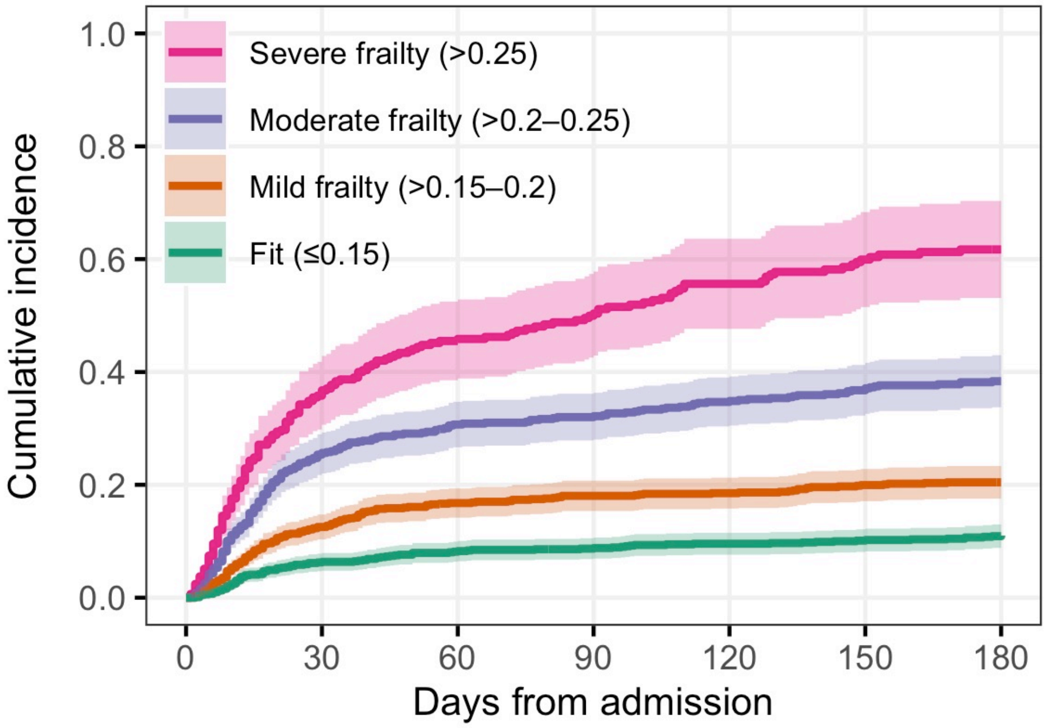

Number at risk

| | | | | | | | |
|---|---|---|---|---|---|---|---|
| Severe frailty | 460 | 319 | 280 | 257 | 238 | 226 | 218 |
| Moderate frailty | 856 | 658 | 616 | 604 | 588 | 571 | 542 |
| Mild frailty | 1061 | 917 | 870 | 848 | 843 | 831 | 799 |
| Fit | 1028 | 958 | 933 | 922 | 911 | 904 | 871 |

**Supplementary Figure 3.** Kaplan-Meier curves for all-cause mortality by categories of the electronic frailty index (n = 3,405)

**Supplementary Table 1.** List of the 48 deficit items for construction of the electronic frailty index

| No. | Deficits | Data source |
|---|---|---|
| **Diagnosis code-based deficits** | | |
| 1 | Anemia | ICD-10 codes |
| 2 | Asthma | ICD-10 codes |
| 3 | Atrial fibrillation | ICD-10 codes |
| 4 | Cancer | ICD-10 codes |
| 5 | Chronic pain | ICD-10 codes |
| 6 | Congestive heart failure | ICD-10 codes |
| 7 | Coronary atherosclerosis and other heart disease | ICD-10 codes |
| 8 | Dementia | ICD-10 codes, MNA |
| 9 | Depression | ICD-10 codes |
| 10 | Diabetes | ICD-10 codes |
| 11 | Dizziness or vertigo | ICD-10 codes |
| 12 | Dyspnea | ICD-10 codes |
| 13 | Fragility fracture | ICD-10 codes |
| 14 | Hypertension | ICD-10 codes, lab values |
| 15 | Hypotension/syncope | ICD-10 codes |
| 16 | Liver disease | ICD-10 codes |
| 17 | Myocardial infarction | ICD-10 codes |
| 18 | Osteoporosis | ICD-10 codes |
| 19 | Parkinsonism and tremor | ICD-10 codes |
| 20 | Peptic ulcer | ICD-10 codes |
| 21 | Peripheral vascular disease | ICD-10 codes |
| 22 | Pulmonary disease | ICD-10 codes |
| 23 | Renal disease | ICD-10 codes |
| 24 | Rheumatoid arthritis or osteoarthritis | ICD-10 codes |
| 25 | Skin ulcer | ICD-10 codes |
| 26 | Stroke or transient ischemic attack | ICD-10 codes |
| 27 | Thyroid disease | ICD-10 codes |
| 28 | Urinary system disease | ICD-10 codes |
| 29 | Valvular disease | ICD-10 codes |
| **Functioning and other health indicators** | | |
| 30 | Activity limitation | Norton, MNA |
| 31 | Cognitive impairment | Norton, Downton |
| 32 | Falls | Downton |
| 33 | Food intake status | Norton, MNA |
| 34 | General condition | Norton |
| 35 | Incontinence | Norton, Barthel |
| 36 | Mobility | Norton |
| 37 | Oral health | ROAG |
| 38 | Sensory impairment | Downton |
| 39 | Weight loss | MNA |
| **Laboratory/anthropometric measures** | | |
| 40 | C-reactive protein | Lab values |
| 41 | Creatinine | Lab values |
| 42 | Glucose | Lab values |
| 43 | Hemoglobin | Lab values |
| 44 | Obesity | Lab values |
| 45 | Potassium | Lab values |
| 46 | Pulse | Lab values |
| 47 | Sodium | Lab values |
| 48 | Underweight | Lab values |

*Note:* The eFI was calculated only if a patient had (i) ≥30 non-missing deficit items and (ii) at least half of the functional and/or laboratory items being non-missing. Norton = Norton scale for pressure ulcer risk assessment; Downton = Downton fall risk assessment scale; MNA = Mini-Nutritional Assessment Scale; Barthel = Barthel scale for activities of daily living; ROAG = Revised Oral Assessment Guide

**Supplementary Table 2.** Sample characteristics by the four study periods (n = 3,405)[a]

| Characteristic | **Period 1:** 3/1/2020 to 31/8/2020 | **Period 2:** 1/9/2020 to 31/1/2021 | **Period 3:** 1/2/2021 to 20/4/2021 | **Period 4:** 21/4/2021 to 31/10/2021 | *p*[b] |
|---|---|---|---|---|---|
| Number of patients, n (%) | 1,193 (35.0) | 1,228 (36.1) | 659 (19.4) | 325 (9.5) | |
| Age, year, mean ± SD | 82.9 ± 8.6 | 82.7 ± 7.9 | 79.8 ± 8.2 | 79.7 ± 9.8 | <0.001 |
| Age category, n (%) | | | | | <0.001 |
| <65 years | 22 (1.8) | 16 (1.3) | 13 (2.0) | 20 (6.2) | |
| 65–74 years | 191 (16.0) | 180 (14.7) | 159 (24.1) | 83 (25.5) | |
| 75–84 years | 437 (36.6) | 507 (41.3) | 302 (45.8) | 111 (34.2) | |
| 85–94 years | 448 (37.6) | 457 (37.2) | 150 (22.8) | 92 (28.3) | |
| ≥95 years | 95 (8.0) | 68 (5.5) | 35 (5.3) | 19 (5.8) | |
| Men, n (%) | 542 (45.4) | 563 (45.8) | 311 (47.2) | 158 (48.6) | 0.71 |
| eFI, median [IQR] | 0.193 [0.150, 0.234] | 0.183 [0.144, 0.227] | 0.163 [0.124, 0.206] | 0.174 [0.136, 0.228] | <0.001 |
| eFI category, n (%) | | | | | <0.001 |
| Fit (≤0.15) | 300 (25.1) | 362 (29.5) | 270 (41.0) | 96 (29.5) | |
| Mild frailty (>0.15–0.2) | 369 (30.9) | 381 (31.0) | 205 (31.1) | 106 (32.6) | |
| Moderate frailty (>0.2–0.25) | 336 (28.2) | 321 (26.1) | 132 (20.0) | 67 (20.6) | |
| Severe frailty (>0.25) | 188 (15.8) | 164 (13.4) | 52 (7.9) | 56 (17.2) | |
| CFS score, median [IQR] | 6.00 [5.00, 7.00] | 5.00 [4.00, 6.00] | 4.00 [3.00, 6.00] | 5.00 [3.00, 6.00] | <0.001 |
| CFS category, n (%) | | | | | <0.001 |
| 1–3 | 61 (5.1) | 127 (10.3) | 99 (15.0) | 38 (11.7) | |
| 4–5 | 183 (15.3) | 246 (20.0) | 126 (19.1) | 53 (16.3) | |
| 6–9 | 311 (26.1) | 311 (25.3) | 100 (15.2) | 47 (14.5) | |
| Missing | 638 (53.5) | 544 (44.3) | 334 (50.7) | 187 (57.5) | |
| HFRS, median [IQR] | 2.60 [1.10, 4.70] | 2.20 [0.60, 4.40] | 1.60 [0.00, 3.80] | 1.80 [0.00, 4.00] | <0.001 |
| HFRS category, n (%) | | | | | 0.003 |
| Low risk (<5) | 910 (76.3) | 967 (78.7) | 544 (82.5) | 265 (81.5) | |
| Intermediate risk (5–15) | 275 (23.1) | 260 (21.2) | 115 (17.5) | 59 (18.2) | |
| High risk (>15) | 8 (0.7) | 1 (0.1) | 0 (0.0) | 1 (0.3) | |
| CCI, median [IQR] | 1 [0, 2] | 1 [0, 2] | 1 [0, 2] | 1 [0, 2] | <0.001 |
| In-hospital mortality, n (%) | 168 (14.1) | 109 (8.9) | 29 (4.4) | 18 (5.5) | <0.001 |
| Discharged to home, n (%) | 747 (62.7) | 751 (61.2) | 471 (71.5) | 220 (67.7) | <0.001 |
| 30-day readmission[c], n (%) | 102 (13.7) | 83 (11.1) | 57 (12.1) | 12 (5.5) | 0.009 |
| Length of stay, day, median [IQR] | 9.02 [6.33, 14.25] | 8.96 [5.93, 13.86] | 7.76 [4.94, 11.81] | 7.87 [5.89, 11.90] | <0.001 |

*Note*: CCI, Charlson Comorbidity Index; CFS, Clinical Frailty Scale; eFI, electronic frailty index; HFRS, Hospital Frailty Risk Score; IQR, interquartile range; SD, standard deviation.

[a] The four study periods correspond to the 1st pandemic wave, 2nd pandemic wave, 3rd pandemic wave, and the time after which most of the patients in our sample were assumed to be fully vaccinated, respectively. Waves of the COVID-19 pandemic were assigned according to statistics of the disease spreading pattern from the Public Health Agency of Sweden.

[b] *P*-values for comparison between study periods, based on ANOVA or Kruskal-Wallis tests for continuous variables, and $\chi^2$ tests for categorical variables

[c] Only patients discharged to home after the first admission were included for analysis of 30-day readmission

**Supplementary Table 3.** Spearman's correlations between frailty and comorbidity measures (n = 1,702)

| *ρ* | CFS | HFRS | CCI | eFI |
|---|---|---|---|---|
| CFS | 1 | | | |
| HFRS | 0.332 | 1 | | |
| CCI | 0.179 | 0.232 | 1 | |
| eFI | 0.464 | 0.365 | 0.419 | 1 |

*Note*: CFS, Clinical Frailty Scale; HFRS, Hospital Frailty Risk Score; CCI, Charlson Comorbidity Index; eFI, electronic frailty index

**Supplementary Table 4.** Logistic regression models for the associations between frailty and comorbidity measures and 30-day readmission (n = 2,189)[a]

| Model | **30-day readmission** | |
|---|---|---|
| | Adjusted OR (95% CI) | AUC |
| **eFI** | | |
| Continuous (per 10% increase) | 1.34 (1.06, 1.68)* | 0.627 |
| Categorical | | |
| Fit (≤0.15) | 1 (Ref.) | 0.623 |
| Mild frailty (>0.15–0.2) | 1.34 (0.96, 1.87) | |
| Moderate frailty (>0.2–0.25) | 1.39 (0.96, 2.00) | |
| Severe frailty (>0.25) | 1.35 (0.82, 2.18) | |
| **CFS (n = 1,065)**[b] | | |
| Continuous (per point increase) | 1.28 (1.12, 1.47)* | 0.659 |
| Categorical | | |
| 1–3 | 1 (Ref.) | 0.649 |
| 4–5 | 1.54 (0.88, 2.83) | |
| 6–9 | 2.41 (1.37, 4.46)* | |
| **HFRS** | | |
| Continuous (per point increase) | 1.03 (0.98, 1.07) | 0.619 |
| Categorical | | |
| Low risk (<5) | 1 (Ref.) | 0.620 |
| Intermediate risk (5–15) | 1.24 (0.88, 1.71) | |
| High risk (>15) | Not estimable | |
| **CCI** | | |
| Continuous (per point increase) | 1.06 (0.97, 1.15) | 0.617 |

*Note*: AUC, area under the receiver operating characteristic curve; CCI, Charlson Comorbidity Index; CFS, Clinical Frailty Scale; CI, confidence interval; eFI, electronic frailty index; HFRS, Hospital Frailty Risk Score; OR, odds ratio

[a] All the listed models were multivariate logistic regression models adjusted for age and sex. Only patients discharged to home after the first admission were included in the 30-day readmission analysis. eFI, CFS, and HFRS were used as both continuous and categorical variables in separate models, while CCI was used as continuous variable only.

[b] Sample size was smaller in analysis of CFS due to missing data

* $p<0.05$

**Supplementary Table 5.** Linear regression models for the associations between frailty and comorbidity measures and length of stay (n = 3,405)[a]

| Model | **Length of stay** | |
|---|---|---|
| | Adjusted $\beta$ (95% CI) | $R^2$ |
| **eFI** | | |
| Continuous (per 10% increase) | 2.28 (1.90, 2.66)* | 6.18% |
| Categorical | | |
| Fit (≤0.15) | 0 (Ref.) | 5.67% |
| Mild frailty (>0.15–0.2) | 1.70 (1.11, 2.29)* | |
| Moderate frailty (>0.2–0.25) | 3.01 (2.38, 3.64)* | |
| Severe frailty (>0.25) | 3.44 (2.68, 4.21)* | |
| **CFS (n = 1,702)**[b] | | |
| Continuous (per point increase) | 0.84 (0.62, 1.07)* | 4.61% |
| Categorical | | |
| 1–3 | 0 (Ref.) | 4.80% |
| 4–5 | 1.84 (0.84, 2.84) | |
| 6–9 | 3.69 (2.70, 4.68)* | |
| **HFRS** | | |
| Continuous (per point increase) | 0.37 (0.30, 0.45)* | 4.96% |
| Categorical | | |
| Low risk (<5) | 0 (Ref.) | 4.14% |
| Intermediate risk (5–15) | 2.31 (1.74, 2.88)* | |
| High risk (>15) | 0.70 (-3.57, 4.97) | |
| **CCI** | | |
| Continuous (per point increase) | 0.39 (0.23, 0.54)* | 3.04% |

*Note*: CCI, Charlson Comorbidity Index; CFS, Clinical Frailty Scale; CI, confidence interval; eFI, electronic frailty index; HFRS, Hospital Frailty Risk Score

[a] All the listed models were multivariate linear regression models adjusted for age and sex. eFI, CFS, and HFRS were used as both continuous and categorical variables in separate models, while CCI was used as continuous variable only.

[b] Sample size was smaller in analysis of CFS due to missing data

* $p$<0.05
